# The Philosophical Perspective in Transculturizing Instrument Tools to Detect Diabetic Foot Ulcer : A Literature Review

**DOI:** 10.1101/2023.12.16.23300092

**Authors:** Sri Wahyuni Awaluddin, Moses Glorino Rumambo Pandin, Nursalam

## Abstract

Diabetes foot ulcer (DFU) is one of the main complications of diabetes mellitus (DM) and one of the main causes of death worldwide from non-communicable diseases (NCDs). The development of an instrument tool for the early diagnosis of diabetic foot is crucial since the findings can be used to prioritize clinical examinations for patients who are at risk. This literature review aimed to examine the philosophical viewpoint on transculturalizing instrument instruments to detect DFU from a variety of study reports published in both domestic and foreign journals. The original research publications published between January 2018 and December 2023 were the focus of the literature search. This study used secondary data, which came from 17 credible journal publications obtained from Science Direct, CINAHL, Pubmed, and SCOPUS databases. The PICOS framework was utilized to assess the papers’ suitability after they were selected. Articles meeting the inclusion requirements would be chosen: the studies should show the outcome of DFU detection or intervention for the outcome, and the population should consist of DM patients with or without DFU, with early DFU detection or intervention for Intervention. The studies could use any kind of research design, including descriptive, cross-sectional, observational, quasi-experimental, randomized controlled trials, and mixed methods, and should be written in English. The result of this literature review showed all the newly developed instrument tools to detect DFU have tested the validity and reliability of content, particularly translation to the local language to meet cultural appropriateness. It is important for researchers working on new DFU detection risk tools to consider including transcultural theory in their assessment instruments for DFU early detection.

## Introduction

Diabetes Mellitus (DM) disease has been known as one of the main global causes of non-communicable diseases (NCDs)-related mortality, which has emerged as a pandemic issue. This disease is characterized by a chronic progressive metabolic disorder presented by hyperglycemia, which is caused by deficiency or resistance to the hormone insulin (Tony I. Oliver; Mesut Mutluoglu., 2023). DM occurs because the pancreas no longer produces insulin, or when the body experiences resistance so that it cannot properly use its insulin. Hyperglycemia, or elevated blood glucose, is a result of the body’s failure to properly make and use insulin. Elevated glucose levels pose a high risk of damage to body tissue and tissue and organ failure in the long term (IDF, 2022).

Various epidemiological studies have reported a trend of increasing incidence and prevalence of DM in various parts of the world every year. A report from the International Diabetes Federation (IDF) in 2023 stated that 537 million persons globally between the ages of 20 and 79 have diabetes. Thus, it is estimated that 1 in 10 people in the world have DM, and 3 out of 4 DM patients come from low-middle-income countries. Adults with DM are predicted to reach up to 643 million in 2030 and 783 million in 2045. Type 2 DM globally accounts for 98% of diagnosed DM patients, although the proportion differs in each country (IDF, 2022).

In both industrialized and developing nations, DM has emerged as a major worldwide health concern. Approximately 11.6% of the US population, or 38.4 million people, have DM (CDC, 2023). In 2021, the Australian Institute of Health and Welfare reported that 1.3 million people suffer from DM, of which almost 1.2 million (4.6%) suffer from Type 2 DM (Reardon et al., 2020). Adults with DM are expected to make up 113 million of the population in Southeast Asia by 2030 and by 2045, that figure is anticipated to have increased by 68% to 151 million. (IDF, 2022). Globally, 642 million people are estimated to have diabetes by 2040 with Type 2 DM accounting for more than 90% of cases. (Harding et al., 2019).

Chronic DM patients with uncontrolled blood sugar levels will face the risk of various complications, both macrovascular and microvascular. Diabetic foot is a chronic consequence of diabetes, that significantly lowers the quality of life for those who have the disease and requires costly treatment (Hnit, Han, and Nicodemus, 2022). Diabetic foot or DFU is caused by peripheral arterial disease (PAD) with sensory neuropathy that occurs in the feet of DM patients (Wang et al., 2022). Peripheral neuropathy causes DM patients to experience sensory loss and ischemia due to peripheral vascular disease and will continue with the occurrence of ulcers in areas of the feet that often experience impact and pressure (McDermott et al., 2022) and end in leg amputation (Clinic, 2022). The following conditions can increase the risk of diabetic foot: sensory, motor, or autonomous neuropathy; peripheral artery occlusive disease (PAOD); restricted joint movement; foot deformities from continuous abnormal pressure (such as wearing inappropriate shoes, having abnormal toes, or being obese); callus, which is a sign of improper distribution of foot pressure; and biopsychosocial variables (e.g., social support deficiency, depression, neglect, and specific ideas about sickness) (McDermott et al., 2022).

DM patients who experience diabetic foot complications (DFU), will result in a decrease in the body’s functional status, infection, longer hospital stays, and further amputation of the lower extremities, as well as death Diabetes patients’ quality of life is significantly impacted by diabetic foot, a chronic problem that requires costly treatment (Hnit et al., 2022). A diabetic patient’s lifetime chance of acquiring a DFU is between 19% and 34%; this risk rises with the patient’s age and level of medical complexity. After ulcers, the recurrence rate is 65% within 3-5 years, which results in rather significant mortality (morbidity). The 5-year death rate is 50–70%, while the lifetime incidence is 20% for lower extremity amputations. According to recent data, the frequency of amputation has increased by up to 50% in some nations overall, particularly among younger populations and racial and ethnic minorities. Late-stage consequences of diabetic foot ulcer (DFU) include amputation and death, and poor diabetes management is significantly linked to these outcomes. There’s a growing disparity in the quality of diabetes care, and current initiatives to enhance care for DFU patients have not led to consistently decreasing rates of amputation. (McDermott et al., 2022).

By taking preventive action, the prevalence of diabetic foot problems can be decreased. According to IDF (2022), a greater focus is needed on preventing diabetic foot, rather than focusing on its treatment, because there are great difficulties in treating and recovering limbs after injury. Prevention and early detection of DFU need to be carried out through a multidisciplinary care approach using guidelines specifically designed to reduce morbidity and treatment gaps associated with DFU (McDermott et al., 2022). For the prevention of diabetic foot, it is necessary to screen DM patients through clinical examination, which produces a risk score for DFU. However, screening activities require time, knowledge, training, and skills from professional health workers, in this case nurses or doctors, to provide effective results. Providing education and screening for the diabetic foot in DM patients is not carried out optimally due to numerous responsibilities that must be carried out by health workers (Sari et al., 2022). For this reason, it is important to develop an instrument for early detection of diabetic foot that is easy and capable of quickly identifying patients who are more susceptible to diabetic foot, where the results can help determine priorities for clinical examination of patients at risk.

Numerous research projects have been undertaken to create tools that can be utilized effectively to prevent diabetic foot in diabetic patients. To explain behavior in preventing DFU, the Health Belief Model appears to be the most popular approach. (Nursalam *et al*., 2020; Sukartini *et al*., 2020). However, another researcher tried to develop a DFU prevention model by integrating psychosocial perspectives, attitudes, intentions, and patient’s coping mechanisms(Pakaya et al., 2020). Evidence-based recommendations for the treatment and prevention of diabetic foot disease have been published by “the International Working Group on Diabetes Foot (IWGDF”) since 1999. (Bus et al., 2023). This instrument has also been used widely in various countries, with several modifications adapted to local conditions (Olarinoye et al., 2021; Vibha et al., 2018; Zantour et al., 2020). Indeed, the success of early detection of diabetic foot depends on various related factors. The newly developed instrument tool for DFU detection risk should be culturally appropriate to enable the patients and their families to apply (Shen, 2015). According to Giger and Davidhizar (1995), a nurse should realize that a patient’s culture can and does influence how he/she is viewed and the care that is provided. This literature review was intended to review the philosophical perspective in transculturizing instrument tools to detect diabetic foot ulcers from various study reports, from national and international publications.

## Design and Method

### Information sources and search strategy

The literature search was undertaken on original research articles published between January 2018 and December 2023. The data used in this study were secondary data, obtained from 25 reputable journal articles from several countries. These articles were obtained from four journal databases: Science Direct, CINAHL, Pubmed, and SCOPUS database. The specific keywords identified by using the “MeSH terms” were “diabetic foot ulcer” OR foot ulcerations OR “diabetic foot” detection AND Diabetes type 2 AND culture.

### Study eligibility and selection criteria

The articles were chosen and their appropriateness was evaluated using the PICOS format. Articles that satisfied the included criteria would be selected: Population (P) should comprise DM patients with or without DFU, with early DFU detection or intervention for Intervention (I), the studies should show the result of Detection or Intervention for DFU for the Ourcome (O), and the studies would be any varies of research design such as Descriptive, Cross-sectional, Observational, Quasy Experimental, RCT and mixed methods. All studies should written in English. Using the exclusion criteria, the research articles were eliminated if: the population of the study was neuropathy patients without DM, the intervention of the study did not discuss DFU interventions, the assessment did not specifically detect the DFU, research articles were published as reviews (literature review, narrative review, scoping review, systematic review, etc), and used non-English language. Then, we reviewed selected articles obtained based on predetermined keywords.

Four hundred and fourteen articles match predetermined keywords that we obtained from Pubmed (n=226), Science Direct (n=143), CINAHL (n=32), and Scopus database (n=13). Following the duplication check, twenty-two articles were excluded, and 202 articles were excluded because they satisfied the exclusion criterion. There were 392 articles left after title identification and abstract screening. Following our retrieval attempt, 102 articles need to be changed, which means that 85 articles are excluded based on the “inclusion and exclusion” criteria. Seventeen papers were chosen for this review based on the eligibility requirements. Figure 1 shows the article selection procedure using the PRISMA flowchart.

Table 1 below lists the features of the studies that were reviewed.

**Figure 1.**
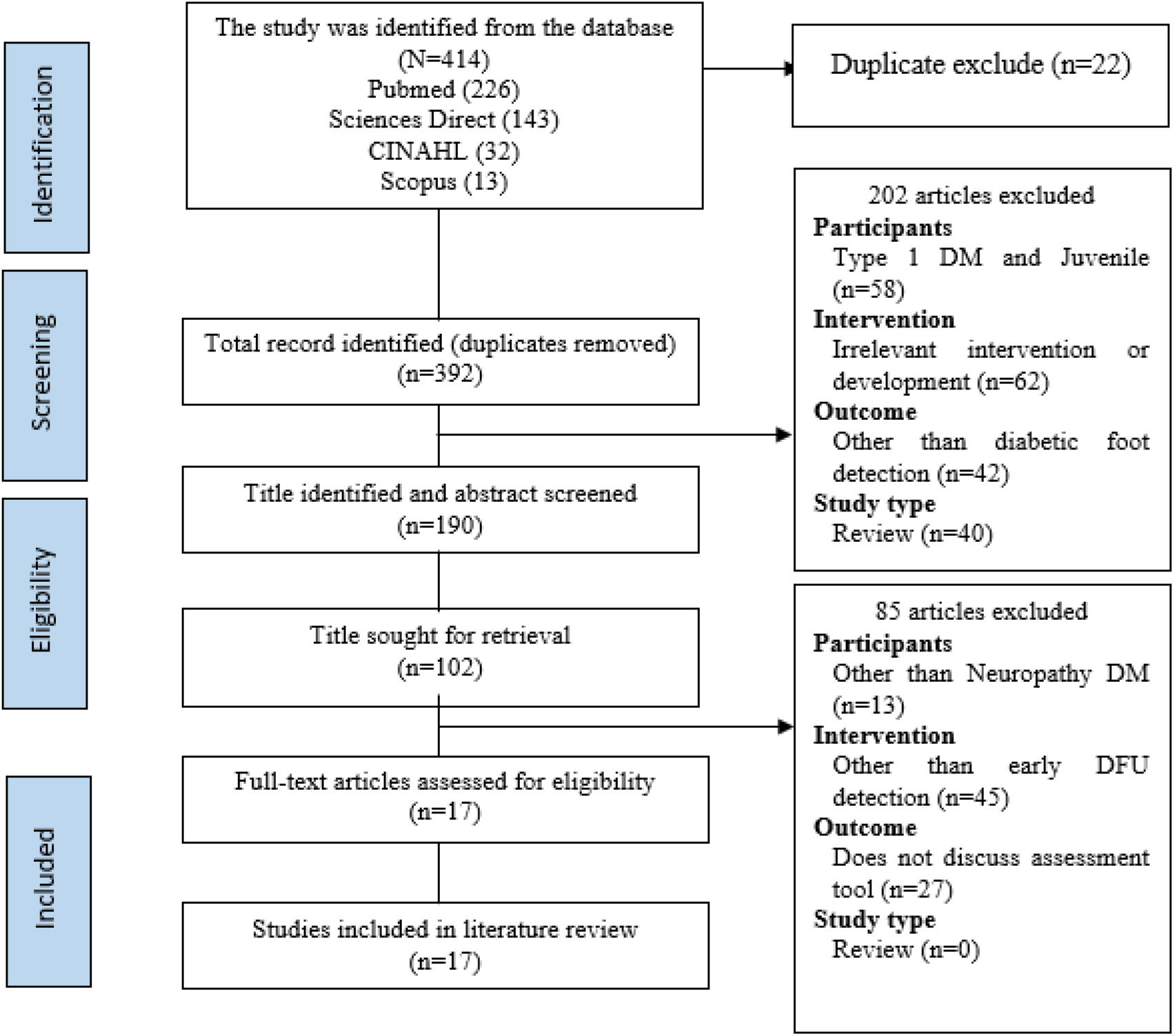
Preferred Reporting Items for Systematic Review and Meta Analysis (PRISMA)

**Table 1.** The Outcome of Article Analysis.

| No | Author | Method | Results |
| --- | --- | --- | --- |
| 1 | Parliani and Phutthikhamin (2020) | **Design:** Descriptive study<br>**Sample:** Nurse at West Kalimantan wound care clinic<br>**Variable:** history of suffering from DM, ulceration history, claudication history, sensory neuropathy, the unusual appearance of the skin, and foot care<br>**Instrument:** Researchers' newly developed DFU risk assessment instrument<br>**Analysis:** "validity, intra-rater, inter-rater, and test-retest reliability" | The instrument has met content validity and reliability.<br>Some input from participating nurses was that the question items regarding ulceration and amputation still needed to be made more specific, several participating nurses suggested further exploration and additional question items.<br>Specialist nurses can understand the instrument, but it is possible that general nurses may not understand some items |
| 2 | Nursalam, Huda and Sukartini (2020) | **Design:** Observational analytics<br>**Sample:** relatives of individuals with type 2 diabetes in a nuclear family<br>**Variable:** family efficacy, family behavior, family behavior, health care facilities, information factor,<br>**Instrument:** questionnaire adjustments to the “NIHM Family Efficacy Scale and the diabetic foot care behavior scale (FCBS)”<br>**Analysis:** “Structural Equation Modeling (SEM) based on variance or Partial Least Squares (PLS)” | • Using social cognitive theory in developing a family foot care model<br>• this strategy has had a significant impact on family behavior.<br>• Families have an impact on efficacy-based foot care; those with high efficacy are crucial in encouraging DFU preventive behavior. |
| 3 | (Sukartini et al., 2020) | **Design:** cross-sectional methodology combined with an explanatory observational design<br>**Sample:** 113 DM patients seeking treatment at the Sidoarjo Regional Hospital polyclinic<br>**Variable:** Age, gender, education, and knowledge are predisposing variables; using health facilities and accessibility are supportive elements; “the role of health workers and family support” are driving factors; attitudes, subjective norms, and self-control are major factors; intention and DFU prevention behavior<br>**Instrument:** Questionnaire<br>**Analysis:** Partial Least Square. | • Knowledge-based predisposing factors, driving factors, and supporting factors have a major impact on the primary factors (behavior attitude, subjective norms, and self-control perception). |
| 4 | Pakaya *et al.* (2020) | **Design:** Cross-sectional<br>**Sample:** 329 DM patients aged 18-85 years in health service facilities.<br>**Variable:** “Knowledge and stress, attitude, intention, coping mechanisms, and foot ulcer prevention”<br>**Instrument:** Questionnaire<br>**Analysis:** SEM-PLS | • All variables contribute indirectly to the prevention of diabetic foot injuries.<br>• The most important direct contribution to the diabetic foot is the patient's coping mechanisms and intentions |
| 5 | Pakaya, Nasrun, Kusnanto, (2020) | **Design:** Cross-sectional<br>**Sample:** 329 DM patients without DFU from 10 health centers<br>**Variable:** Social support (family and friends), perceived control, self-efficacy, intention to diet, taking medication, physical activity, checking blood sugar/legs<br>**Instrument:** Questionnaire<br>**Analysis:** SEM-PLS | • Social support contributes directly to the desire (intention) to prevent DFU |
| 6 | Ma *et al.* (2022) | **Design:** Descriptive study<br>**Sample:** 208 DM patients<br>**Variable:** “Quality of Life (diabetic foot ulcer scale), Physical Functioning, Role-Physical, Bodily Pain, General Health, Validity, Social Functioning, | • It will be a trustworthy tool to assess the quality of life of Chinese patients with diabetic foot ulcers because the adaption and validation of the Diabetic Foot Ulcers Scale- |
|  |  | Role-Emotional, Mental Health, and Reported Health Transition”.<br>**Instrument:** “The Diabetic Foot Ulcer Scale-Short Form (DFS-SF)” in Chinese version<br>**Analysis:** “Cronbach's $\alpha$ coefficient, split-half reliability, and test-retest reliability were used to evaluate the reliability, while content validity, structural validity, and criteria validity techniques” were used to validate the scale | Short Form were credible.<br>• The use of Brislin's translation model needs to be applied in the back translation tool |
| 7 | Luo *et al.*, (2023) | **Design:** Descriptive cross-sectional study<br>**Sample:** Patients with DM<br>**Variable:** “Validity and Reliability of the Chinese version of DFUA”<br>**Instrument:** “Diabetic foot ulcer assessment scale (DFUAS) and Bates-Jensen wound assessment tool (BWAT)”<br>**Analysis:** “The Pearson correlation and Bland–Atman plots were used to analyzing the consistency between the DFUAS and the BWAT”. | • “The Diabetic Foot Ulcers Scale-Short Form” adaptation and validation for use in China were dependable, and it will be a trustworthy tool for assessing the QoL of Chinese patients with DFU. |
| 8 | Al-Busaidi, Abdulhadi, and Coppell,(2020) | **Design:** A cross-sectional survey<br>**Sample:** 353 consecutive Omanis with diabetes who are 20 years of age or older<br>**Variable:** “Clinico-demographic characteristics, patient-reported foot complications, and foot self-care practices”<br>**Instrument:** a recently created and trialed survey<br>**Analysis:** Descriptive statistics | • “The Diabetic Foot Disease and Foot Care Questionnaire (DFDFC-Q)” was chosen as the new questionnaire to be created since<br>• Oman does not already have a survey instrument that is culturally appropriate for measuring independent foot care. |
| 9 | Tavassolmand (*et al.*, 2023) | **Design:** Descriptive study<br>**S:** 262 DM patients<br>**Variable:** Using the “content validity index (CVI), criterion validity, content validity ratio (CVR), and Validity of construction”<br>**Instrument:** Short form of the DFU scale in Persian<br>**Analysis:** “Cronbach's alpha, intraclass correlation coefficient (ICC), and Spearman correlation” | • The QoL, is a widely utilized outcome in intervention evaluation.<br>• The DFS-SF offers sufficient information to evaluate the QoL of DFU patients.<br>• The Persian DFS-SF is valid and reliable; as a result, it can be used to assess patients' QoL in clinical and research contexts. |
| 10 | Aksoy, Büyükbayram and Özüdoğru (2023) | **Design:** Study that is cross-sectional, descriptive, and methodological<br>**Sample:** 193 people with diabetes who attend the internal medicine outpatient clinics<br>**Variable:** disease history according to sociodemographic characteristics | • “The Turkish version of the Foot Posture Index (FPI-6)” has been found to have excellent intra-, inter-, and internal consistency. Also, mediocre relationships with “the Foot Function Index (FFI)” and the |
|  |  | (disease type, diabetes medication, family history, foot care education, foot wound condition).<br>**Instrument:** “The Diabetic Foot Self-Care Questionnaire (DFSQUMA) from the University of Malaga”<br>**Analysis:** “The Kaiser-Meyer Olkin (KMO) and Bartlett x2 tests. (KMO)” | American Orthopedic Foot and Ankle Society (AOFAS)” were found.<br>• “ The Diabetic Foot Self-Care Questionnaire, the Self-care of Hypertension Inventory (SC-HI), and the Diabetes Foot Self-Care Behavior Scale (DFSBS)” were found to have strong psychometric qualities in their Turkish versions. These qualities included strong internal consistency, a high index of content validity, and high item-level content validity. |
| 11 | Abrar *et al.* (2020) | **Design:** The cross-sectional inquiry was conducted in three stages. In the first phase, the Delphi study's consensus was used to create video educational programs on “diabetic foot care”. The second phase entailed creating and approving the instructional video about “diabetic foot care” by the researchers. Following their viewing of the film, the third stage involved the researchers evaluating the level of knowledge the diabetic patients from Makassarese and Buginese possessed regarding foot care.<br>**Sample:** 20 DM Individuals from both the Makassarese and Buginese ethnic groups<br>**Variable:** The patient’s knowledge and an instructional film about “diabetic foot care” in the local languages of Makassarese and Buginese are the variables.<br>**Instrument:** Online Delphi application, assessment of video content, patients’ pre and post-test questionnaire<br>**Analysis:** Descriptive statistics and the Wilcoxon test | • The Delphi survey yielded five themes: identifying pre-ulcer symptoms, cleaning feet, trimming nails, donning socks, and inspecting shoes.<br>• The assessment of content validity suggests incorporating these things into instructional videos produced in local languages.<br>• The evaluation conducted in the community revealed a noteworthy ($p = 0.001$) increase in diabetic people with DM who were also at risk of developing DFU understanding of foot care. |
| 12 | Musdiaman<sup>1</sup> *et al.*, (2020) | **Design:** descriptive research methods.<br>**Sample:** Forty families with prior research education<br>**Instrument:** a questionnaire with ten question items; Variable: family knowledge in identifying the risk of DFU<br>**Analysis:** Descriptive frequency | • Family knowledge about early detection of DFU is better at young ages than at old ages<br>• Family knowledge (25% vs 2.5%), and especially housewives have good knowledge skills |
| 13 | Şahin and Cingil, (2020a) | **Design:** Descriptive with a component of clinical evaluation<br>**Sample:** 246 individuals with type 2 DM who are treated at a family health | • The DFSBS scores showed a significant difference “($p < 0.05$)” according to the patients' gender and whether they lived in |
|  |  | center in Konya, Turkey.<br>**Variable:** foot wound risk, illness acceptance<br>**Instrument:** psychometric questionnaires:<br>1. "Socio-demographic questionnaire"<br>2. "The Michigan Neuropathy Screening Instrument Questionnaire (MNSI-Q)"<br>3. "Diabetes Foot Self-Care Behavior Scale (DFSBS)"<br>4. "Acceptance of Illness Scale (AIS)"<br>5. "Omaha System Problem Classification Scheme, which includes the pain, skin, and circulation subscales [Omaha System of Total Number of Signs/Symptoms (OTNS)]"<br>**Analysis:** hierarchical regression analysis | an urban or rural region.<br>• The participants' gender, level of education, social security status, domicile, and financial situation were found to significantly differ from one another in terms of the AIS score ($p < 0.05$).<br>• Regression analysis results showed that gender, the number of monthly physician control visits, and "foot care training" were predictive factors for the DFSBS score.<br>• The AIS and OTNS scores were used to make further predictions for the MNSI-Q score outcomes anticipated the AIS rating. |
| 14 | Yılmaz Karadağ *et al.* (2019) | **Design:** cross-sectional<br>**Sample:** 1030 patients in Turkey between November 2017 and February 2018<br>**Variables:** include age, sex, employment position, educational attainment, socioeconomic situation, and the patients' clinical characteristics and level of awareness.<br>**Instrument:** A questionnaire concerning foot care knowledge and practice;<br>**Analysis:** using the Shapiro-Wilk, Mann Whitney, Kruskal Wallis, and Chi-Square tests | • It is evident from the patients' intermediate mean scores for knowledge and practice that there is still a significant need for patient education on this topic.<br>• Patients with DM were also observed to practice "foot care" to be significantly impacted by factors such as disease duration, education level, and awareness of the significance of this practice. |
| 15 | Vibha *et al.* (2018) | **Design:** Cross-Sectional Study<br>**Sample:** 620 DM patients in rural areas of India<br>**Variable:** Sociodemographics, lifestyle, diabetic foot syndrome<br>**Instrument:** The Michigan Neuropathy Screening Instrument, Ankle-brachial index(ABI), the IWGDF classification system.<br>**Analysis:** Univariate and multivariate regression analysis | Diabetic Foot Syndrome affects a significant portion of the population, it is necessary to carry out diabetic foot screening from the time the patient is diagnosed with DM, integrated with regular health education through training for health workers in primary care facilities |
| 16 | Olarinoye *et al.*, (2021) | **Design:** Cross-sectional study<br>**Sample:** 151 patients with DM were being treated at a teaching hospital in Nigeria.<br>**Variables:** diabetes, foot care, sociodemographics, "dermatological changes, musculoskeletal abnormalities, neurological issues, and vascular<br>problems.”<br>**Instrument:** Questionnaire; IWGDF Risk Classification System<br>A: The Chi-square test, Pearson’s correlation | • The majority of diabetic individuals who experience foot ulcerations have a terrible quality of life, and many of the factors that lead to this can be changed or avoided. |
| 17 | Zantour *et al.*, (2020) | **Design:** Cross-sectional study<br>**Sample:** Over seven months, a random selection process was used to choose patients from the Tahar Sfar Hospital in Mahdia's diabetes-endocrinology outpatient department.<br>**Variable:** “PSN, PAD, deformity, and patient’s history of amputation and/or ulceration”<br>**Instrument:** Questionnaire, along with clinical examination using IWGD diabetic foot classification<br>**Analysis:** The T student's test and “Pearson's Chi2 test” | • Educating diabetics and routinely inspecting feet for lesions;<br>• Paying special attention to patients with retinopathy and low school level or hyperkeratosis. |

## Results and Discussion

Methodologies for conducting research are based on certain methods or methodologies, as well as paradigms and philosophical assumptions (Faith Alele & Malau-Aduli, 2023). Researchers’ methodologies are influenced by their worldviews, which are made up of their philosophical assumptions and notions on the nature of reality and the means of understanding it (Guba EG, Lincoln YS, 1994 in Faith Alele and Malau-Aduli, 2023). Transcultural nursing, which was formed 40 years ago as a formal field of study and practice was aimed to deliver care that is culturally appropriate. Global cultures and comparative cultural nursing, health, and caregiving paradigms are the main subjects of Transcultural nursing. The emphasis of this nursing discipline is on incorporating transcultural and international content into instruction. Examples of subjects covered in the courses are international health issues and organizations, nursing in different nations, and cultural differences (Murphy et al., 2006). This section aims to examine the philosophical viewpoint of researchers using diverse study reports while transculturizing instruments to identify diabetic foot ulcers.

Amongst the 17 articles reviewed, we found that the researchers used various methods or designs to investigate to risk of DFU to discover the best instrument for DFU detection risk. Some researchers used descriptive (Parliani and Phutthikhamin, 2020; Musdiaman *et al*., 2020; Şahin and Cingil, 2020b; Ma *et al*., 2022; Tavassolmand *et al*., 2023) and observational analytic (Nursalam et al., 2020) for their studies. The other researchers used cross-sectional “(Abrar et al., 2020a; Aksoy et al., 2023; Al-Busaidi et al., 2020; Luo et al., 2023; Olarinoye et al., 2021; Pakaya, Nasrun, Kusnanto, 2020; Pakaya et al., 2020; Sukartini et al., 2020; Vibha et al., 2018; Yılmaz Karadağ et al., 2019; Zantour et al., 2020)” to test the new developed DFU detection risk that locally adapted for cultural appropriateness.

The numerous research designs employed in the studies can be interpreted from a philosophical standpoint as proof that the researchers’ worldviews— their approaches are influenced by their philosophical assumptions and notions about the nature of the universe and how it might be comprehended Methodology in the context of a certain research paradigm refers to the strategy or plan of action that directs the choice and implementation of particular techniques. The word “methodology” describes the strategies, techniques, and plans used in a carefully thought-out research project to obtain answers (Faith Alele & Malau-Aduli, 2023).

### 1. Ontological study of transculturizing instrument tools to detect diabetic foot ulcers

The definition of ontology is the accurate representation of reality as an entity or entities, or the nature of reality. Its main focus is on the presumptions researchers make to accept data as true (Faith Alele & Malau-Aduli, 2023). The understanding of the existence and ontological character of the detection tool to be produced is part of the ontology component of the study topic “transculturizing instrument tools to detect diabetic foot ulcers.” Ontology in this sense refers to the type of categories or nature that support the existence of the detecting tool. The ontological idea underlying the detection tool must be thoroughly understood for this research to take into account factors like the entities (clinical data, clinical symptoms, etc.) and their relationships as well as how the entities are classified according to the pertinent specific ontology. Research on diabetic foot ulcers (DFU) emphasizes the use of instruments for wound identification and evaluation to maximize the healing of problems.

A modification of the IWGDF’s early detection instrument for diabetic feet, along with additional information about a history of diabetic foot disease, ulceration, claudication, sensory neuropathy, abnormal skin appearance, and foot care, has been developed by Parliani and Phutthikhamin (2020). This instrument has been tested on specialist wound nurses at the West Kalimantan wound care clinic and has met validity and reliability. Several other researchers have also developed diabetic foot assessment instruments by modifying them according to the needs of their countries. A combination of several instruments to assess the risk of diabetic feet was carried out by Şahin & Cingil, (2020) in Turkey, by combining instruments from “the Problem Classification of the OMAHA System (OTNS), the Diabetes Foot Self-Care Behavior Scale (DFSBS), the Acceptance of Illness Scale (AIS), and the Michigan Neuropathy Screening Instrument Questionnaire (MNSI-Q)”.

### 2. Epistemological study of transculturizing instrument tools to detect diabetic foot ulcers

Understanding how to gather information about the detection tool that is to be produced is part of the epistemological component of the study topic “transculturizing instrument tools to detect diabetic foot ulcers.” The term “epistemology” in this sense refers to the techniques employed to learn about the detection instrument, such as the testing procedures, the tool’s validity, and its dependability. To guarantee the accuracy and dependability of detection results, studies about diabetic foot ulcers (DFU) emphasize the significance of testing and verifying detection techniques.

The need for an instrument that is culturally appropriate is something that is considered important to facilitate the achievement of early detection of diabetic foot. Therefore, many researchers have attempted to modify diabetic foot detection instruments so that they meet local cultural suitability. “The Diabetic Foot Ulcer Assessment Scale (DFUAS)” and “the Chinese version of the Bates-Jensen wound assessment tool (BWAT)” were combined to create an instrument that Luo *et al*., (2023) assessed, and it was successful in meeting the standards for validity and reliability. Likewise, the Iranian researchers, Tavassolmand *et al*. (2023), have confirmed the “validity and reliability” of the Persian-language version of “the Diabetic Foot Ulcers Scale Short Form (DFS-SF)” instrument. To address the lack of a survey instrument that was culturally appropriate for the local community in Oman, Al-Busaidi, Abdulhadi, and Coppell (2020) created “the Diabetic Foot Disease and Foot Care Questionnaire (DFDC-Q)”. The instrument from Al-Busaidi et al. (2020) assesses patient clinical and demographic information, reports of foot problems, and patient “self-foot care”. A study in Makassar, Indonesia, using the traditional languages of Makassarese and Buginese to determine whether the patient’s level of knowledge had changed after watching the instructional video on” diabetic foot care” (Abrar et al., 2020b). An expert panel evaluated the video’s content validity in three stages of the study: a trial conducted in a community setting with patients who were diagnosed with DM, spoke the local languages, and were at risk of developing DFU; also, a Delphi research was conducted with wound-care specialist nurses to develop the video content. The evaluation items in the study were observing pre-ulcer indications, cleaning feet, trimming toenails, donning socks, and inspecting footwear are among the five themes that arose from the Delphi survey. It was suggested by the content validity evaluation that these items be written in traditional languages for use in video education. Patients with DM at risk of DFU had a notable (p = 0.001) increase in foot care knowledge, according to examinations conducted in a community setting.

### 3. Axiological study of transculturizing instrument tools to detect diabetic foot ulcers

The axiology component of the research topic “developing instrument tools to detect diabetic foot ulcers” comprises the values associated with the tool’s creation. Axiology, as used here, refers to the moral principles—such as justice, humanism, and ethics—that guide the creation of instruments. Developing the DFU detection tool to help diagnose diabetic foot ulcers is the aim of this research. The DFU detection tool that is developed must respect the patients’ humanity who are affected by diabetic foot ulcers and follow the strictest ethical and legal guidelines when being used. Additionally, when using the built detection tool, consideration must be given to values like validity, accuracy, and dependability.

Some studies reported good psychometric properties along with high item-level content “validity, and good internal consistency of the translated and adapted version” of their DFU detection risk instrument tools “(Aksoy et al., 2023; Al-Busaidi et al., 2020; Luo et al., 2023; Ma et al., 2022; Şahin & Cingil, 2020b; Tavassolmand et al., 2023; Vibha et al., 2018)”. Based on these results, it can be concluded that integrating transcultural theory into an assessment tool for DFU early detection should be taken into account for those researchers who are attempting to develop a new DFU detection risk tool.

## Conclusions

A diabetic foot detection instrument must be culturally and linguistically adjusted to be considered transcultural. This procedure can enhance the tool’s efficacy and accuracy in identifying diabetic foot problems in a range of demographics. Developing a diabetic foot detection instrument that integrates transcultural theory, the researchers attempt the following procedures :

1. Determine the language and cultural differences. Recognize the variations in the target groups’ languages, cultures, and customs. This will assist in customizing the tool to their requirements and tastes.
2. Content adaptation. Make changes to the tool’s questions, directions, and examples to take into account the linguistic and cultural variances noted in Step 1. This might be combining regional lingo and ideas or translating the tool into other languages.
3. Examine the modified tool. To determine the modified tool’s correctness and efficacy, conduct a pilot test on a subset of the intended audience. Invite users to provide feedback, then act upon their suggestions.
4. Train medical professionals: Educate medical professionals—such as nurses—on the usage of the modified tool. By doing so, they can make sure they can correctly identify diabetic foot issues and use the tool in their practice
5. Evaluate the effectiveness and efficiency. Assess the effectiveness and efficiency of the adapted tool by comparing it to the original version or other detection methods. This can help determine if the adaptation process has improved the tool’s performance.

## Data Availability

All data produced in the present work are contained in the manuscript

## Conflict of Interest

The authors have no conflict of interest.

## Acknowledgment

Our gratitude to Universitas Airlangga and everyone who took part in this literature review. Funding organizations from the governmental, private, or nonprofit sectors did not provide a specific grant for this literature review.

## Notes

### Competing Interest Statement

The authors have declared no competing interest.

### Funding Statement

This study did not receive any funding

## References

Abrar, E. A., Yusuf, S., Sjattar, E. L., & Rachmawaty, R. (2020a). Development and evaluation of educational videos of diabetic foot care in traditional languages to enhance knowledge of patients diagnosed with diabetes and risk for diabetic foot ulcers. Primary Care Diabetes, 14(2), 104–110. 10.1016/j.pcd.2019.06.005

Abrar, E. A., Yusuf, S., Sjattar, E. L., & Rachmawaty, R. (2020b). Development and evaluation of educational videos of diabetic foot care in traditional languages to enhance knowledge of patients diagnosed with diabetes and risk for diabetic foot ulcers. Primary Care Diabetes, 14(2), 104–110. 10.1016/j.pcd.2019.06.005

Aksoy, M., Büyükbayram, Z., & Özüdoğru, O. (2023). Reliability and validity of the Diabetic foot self-care questionnaire in Turkish patients. Primary Care Diabetes, June. 10.1016/j.pcd.2023.06.005

Al-Busaidi, I. S., Abdulhadi, N. N., & Coppell, K. J. (2020). Development and pilot testing of a diabetes foot care and complications questionnaire for adults with diabetes in Oman: The diabetic foot disease and foot care questionnaire. Oman Medical Journal, 35(4), 1–8. 10.5001/omj.2020.65

Bus, S. A., Armstrong, D. G., Crews, R. T., Gooday, C., Jarl, G., Kirketerp-Moller, K., Viswanathan, V., & Lazzarini, P. A. (2023). Guidelines on offloading foot ulcers in persons with diabetes (IWGDF 2023 update). Diabetes/Metabolism Research and Reviews. 10.1002/dmrr.3647

CDC. (2023). National Diabetes Statistics Report Estimates of Diabetes and Its Burden in the United States.

Clinic, M. (2022). Amputation and diabetes: How to protect your feetitle. https://www.mayoclinic.org/diseases-conditions/diabetes/in-depth/amputation-and-diabetes/art-20048262

Faith Alele, & Malau-Aduli, B. (2023). An Introduction to Research Methods for Undergraduate Health Profession Students (J. C. University (ed.); 1st edition). James Cook University. https://jcu.pressbooks.pub/intro-res-methods-health/

Giger, J. N., & Davidhizar, R. E. (1995). Transcultural Nursing, Assessment and Intervention (N. D. Como & L. Sparks (eds.); 2nd Editio). Mosby-Year Book, Inc.

Harding, J. L., Pavkov, M. E., Magliano, D. J., Shaw, J. E., & Gregg, E. W. (2019). Global trends in diabetes complications: a review of current evidence. Diabetologia, 62(1), 3–16. 10.1007/s00125-018-4711-2

Hnit, M. W., Han, T. M., & Nicodemus, L. (2022). Accuracy and Cost-effectiveness of the Diabetic Foot Screen Proforma in Detection of Diabetic Peripheral Neuropathy in Myanmar. Journal of the ASEAN Federation of Endocrine Societies, 37(1). 10.15605/jafes.037.01.06

IDF. (2022). IDF Diabetes Atlas. https://diabetesatlas.org/#:~:text=Diabetes xaround the world in 2021%3A,- and middle-income countries.

Luo, Y. X., Mai, L. F., Liu, X. Z., & Yang, C. (2023). Validity and reliability of the Chinese version of the new diabetic foot ulcer assessment scale. International Wound Journal, 20(9), 3724–3730. 10.1111/iwj.14266

Ma, L., Ma, W., Lin, S., Li, Y., & Ran, X. (2022). Adaptation and Validation of the Diabetic Foot Ulcer Scale-Short Form Scale for Chinese Diabetic Foot Ulcers Individuals. International Journal of Environmental Research and Public Health, 19(21). 10.3390/ijerph192114568

McDermott, K., Fang, M., Boulton, A. J. M., Selvin, E., & Hicks, C. W. (2022). Etiology, Epidemiology, and Disparities in the Burden of Diabetic Foot Ulcers. Diabetes Care, 46(1), 209–211. 10.2337/dci22-0043

Murphy, S. C., Edu, H., & Librarian, A. (2006). Mapping the literature of transcultural nursing*. In J Med Libr Assoc (Vol. 94, Issue 2). https://www.tcns.org

Musdiaman1, S., Yusuf2, S., Afelya3, T. I., Hidayah, N., Studi, P., Keperawatan, I., & Keperawatan, F. (2020). EVALUATION OF FAMILY KNOWLEDGE IN DETECTING RISK OF DIABETES FOOT ULCER IN PUBLIC HEALTH CENTER. In Indonesian Contemporary Nursing Journal (Vol. 4, Issue 2).

Nursalam, N., Huda, N., & Sukartini, T. (2020). Development of efficacy-based foot care by family models to family behavior in prevention of diabetic foot ulcer. Systematic Reviews in Pharmacy, 11(7), 240–245. 10.31838/srp.2020.7.38

Olarinoye, J., Bello, A., Ogunkeyede, S., Aderibigbe, A., Olagbaye, B., & Wahab, K. (2021). Risk assessment for foot ulceration in a Nigerian diabetic population attending University of Ilorin Teaching Hospital, Ilorin. Libyan International Medical University Journal, 06(02), 81–90. 10.4103/liuj.liuj_83_21

Pakaya, Nasrun, Kusnanto, H. B. N. & R. S. T. (2020). Intention of Diabetic Foot Ulcer Prevention Model Based on Social Support and Personal Agency Perspectives. Indian Journal of Public Health, 11(1), 1–8.

Pakaya, N., Kusnanto Notobroto, H. B., & Triyoga, R. S. (2020). The development of a diabetic foot ulcer prevention model based on psychosocial perspectives, attitudes, intentions, and coping mechanisms. In Indian Journal of Public Health Research and Development (Vol. 11, Issue 3, pp. 2215–2221). https://www.embase.com/search/results?subaction=viewrecord&id=L2004452611&from=export

Parliani, & Phutthikhamin, N. (2020). Cultural Appropriateness of the Risk Assessment Tool for Diabetic Foot Ulcer and Its Psychometric Properties Among Diabetes Mellitus Patients. American Journal of Humanities and Social Sciences Research (AJHSSR), 4(9), 113–118.

Reardon, R., Simring, D., Kim, B., Mortensen, J., Williams, D., & Leslie, A. (2020). The_diabetic_foot_ulcer_Australia. Australian Journal of General Practice, 49(5), 250–255.

Şahin, S., & Cingil, D. (2020a). Evaluation of the relationship among foot wound risk, foot self-care behaviors, and illness acceptance in patients with type 2 diabetes mellitus. Primary Care Diabetes, 14(5), 469–475. 10.1016/j.pcd.2020.02.005

Şahin, S., & Cingil, D. (2020b). Evaluation of the relationship among foot wound risk, foot self-care behaviors, and illness acceptance in patients with type 2 diabetes mellitus. Primary Care Diabetes, 14(5), 469–475. 10.1016/j.pcd.2020.02.005

Sari, Y., Yusuf, S., Haryanto Kusumawardani, L. H., Sumeru, A., Sutrisna, E., & Saryono. (2022). The cultural beliefs and practices of diabetes self-management in Javanese diabetic patients: An ethnographic study. Heliyon, 8(2). 10.1016/j.heliyon.2022.e08873

Shen, Z. (2015). Cultural Competence Models and Cultural Competence Assessment Instruments in Nursing: A Literature Review. In Journal of Transcultural Nursing (Vol. 26, Issue 3, pp. 308–321). 10.1177/1043659614524790

Sukartini, T., Theresia Dee, T. M., Probowati, R., & Arifin, H. (2020). Behavior model for diabetic ulcer prevention. Journal of Diabetes and Metabolic Disorders, 19(1), 135–143. 10.1007/s40200-019-00484-1

Tavassolmand, S. S., Montazeri, A., Madadizadeh, F., Dehghan, H. R., Ranjbar, M., & Ameri, H. (2023). Translation and validation of the Persian version of diabetic foot ulcer scaleshort form (DFS-SF). International Wound Journal, 20(3), 822–830. 10.1111/iwj.13929

Tony I. Oliver; Mesut Mutluoglu. (2023). Diabetic Foot Ulcer. StatPearls [Internet]. Treasure Island (FL): StatPearls Publishing. https://www.ncbi.nlm.nih.gov/books/NBK537328/

Vibha, S. P., Kulkarni, M. M., Kirthinath Ballala, A. B., Kamath, A., & Maiya, G. A. (2018). Community based study to assess the prevalence of diabetic foot syndrome and associated risk factors among people with diabetes mellitus. BMC Endocrine Disorders, 18(1), 1–9. 10.1186/s12902-018-0270-2

Wang, X., Yuan, C.-X., Xu, B., & Yu, Z. (2022). Diabetic foot ulcers: Classification, risk factors, and management. World Journal of Diabetes, 13(12), 1049–1065. 10.4239/wjd.v13.i12.1049

Yılmaz Karadağ, F., Saltoğlu, N., Ak, Ö., Çınar Aydın, G., Şenbayrak, S., Erol, S., Mıstanoğlu Özatağ, D., Kadanalı, A., Küçükardalı, Y., Çomoğlu, Ş., Yörük, G., Akkoyunlu, Y., Meriç Koç, M., & Altunçekiç Yıldırım, A. (2019). Foot self-care in diabetes mellitus: Evaluation of patient awareness. Primary Care Diabetes, 13(6), 515–520. 10.1016/j.pcd.2019.06.003

Zantour, B., Bouchareb, S., El Ati, Z., Boubaker, F., Alaya, W., Kossomtini, W., & Sfar, M. H. (2020). Risk assessment for foot ulcers among Tunisian subjects with diabetes: A cross-sectional outpatient study. BMC Endocrine Disorders, 20(1). 10.1186/s12902-020-00608-2

